# Improving Veteran Access to Integrated Management of Back Pain (AIM-Back): Updated Statistical Analysis Plan for this Embedded Pragmatic, Cluster Randomized

**DOI:** 10.1101/2023.12.22.23300382

**Authors:** Cynthia J. Coffman, Rebecca North, Steven Z. George, Susan N. Hastings

## Abstract

The Improving Veteran Access to Integrated Management of Back Pain (AIM-Back) pragmatic, embedded, cluster-randomized trial is ongoing with enrollment starting in February 2020 and projected to end in first quarter of 2024. The 3 -month follow up rate of primary outcome data collected as part of an AIM-Back clinical follow-up visit in the electronic health record has been lower than anticipated. At the recommendation of AIM-Back monitoring partners an updated statistical analysis plan was generated. The updated analysis plan allows for use of survey data to augment the follow up rate for the primary outcomes collected in the electronic health record. This updated statistical analysis plan was created and approved prior to completing enrollment and described in this paper.

## Background

The Improving Veteran Access to Integrated Management of Back Pain (AIM-Back) pragmatic, embedded, cluster-randomized trial is ongoing with key elements of the study design, care pathways, primary outcomes and data analysis included in the published protocol paper.[1] AIM-Back was prospectively registered at www.clinicaltrials.gov (NCT04411420), began enrollment in February 2020, and is expected to continue enrollment into the first quarter of 2024. In response to a recommendation from our Data Safety Monitoring Board and in collaboration with study sponsor (National Center for Complementary and Integrative Health) and the NIH/DoD/VA Pain Management Collaboratory Biostatistics and Study Design Work Group we are updating our analysis plan prior to completing enrollment. The primary motivation for updating our statistical analysis plan is that we have observed a much lower than expected completion of 3-month follow-up where data is collected in the electronic health record (EHR) during AIM-back clinical visits. The updated statistical analysis plan was approved by our monitoring partners on December 18, 2023. Therefore, the purpose of this paper is to provide the rationale and justification for the updated data analysis plan, as well as to fully describe the updated analysis plan. The updated statistical analysis plan is only for our originally planned primary analyses and does not involve a change in the primary outcomes originally described in the protocol paper.

### Ongoing Trial Progress Informing Updated Analysis Plan

The original power calculations for the this EHR sample were done assuming an 80% follow-up rate at 3-months, with 16 clinics enrolling n=105 at each clinic at baseline to have 84 per clinic at 3-months. The effect size difference we are powered to detect assuming an ICC of 0.01 is 0.30 and for an ICC of 0.05 is 0.50. As of the end of September 2023, the overall 3-month EHR completion rate is 51% (52% in the sequenced care pathway (SCP) arm; 49% in pain navigator pathway (PNP) arm). With a 40% completion rate, n=42 at 3-months at each site with all the same assumptions as original power calculation, the effect size difference we can detect is 0.37 for an ICC of 0.01 and 0.55 for an ICC of 0.05. As the ICC is a nuisance parameter[2], we have calculated interim estimates of the ICC for our co-primary outcomes, PROMIS pain interference and physical function, at 3-months adjusted for baseline PROMIS pain interference and physical function, respectively. These calculations indicated they are comparable to our range for the original power calculations on the lower end of the range. To estimate the adjusted ICC, we fit a mixed model with the 3-month PROMIS outcome measure and adjusted for baseline PROMIS measure fitting a random effect for clinic, and used the covariance estimates for calculating the ICC. The estimated adjusted ICCs for the primary outcomes in the EHR sample and bootstrapped 95% confidence intervals are 0.014 with 95% CI of [0.00,0.046] for PROMIS physical function and 0.001 with 95% CI of [0.00,0.028] for PROMIS pain interference. The lower bound for the bootstrapped confidence interval for the ICC for both physical function and pain interference is 0.

During the trial we have enrolled and randomized 19 clinics; however, 2 clinics in block 2 withdrew from delivering the AIM-Back program after 8 months. Of those clinics that withdrew, 1 Veteran had enrolled with 3 referrals at one of the clinics and 5 Veterans enrolled with 9 referrals at the second clinic. For participating clinics the enrollment goal was to have clinics be in the 65-130 Veterans range with enrollment capped at n=130 to reduce variability in cluster sizes across clinics. As of the end of September 2023, n=1752 Veterans have enrolled with 12 clinics having met or exceeded the n=105 goal; 3 clinics have met minimum enrollment goals of n=65. We plan to complete enrollment in the first quarter of 2024 and project that 13-15 clinics will meet or exceed the n=105 enrollment goal. Current projections are that 2 clinics may be under the n=65 minimum goal.

### Additional Data Source

We have collected the primary PROMIS outcomes for pain interference and physical function in the EHR. As described in our protocol paper[1], Veterans that are referred to the AIM-Back program by providers can also agree to be contacted for a survey study. These Veterans provide consent to participate, and are administered surveys that include the primary PROMIS measure outcomes and additional measures at baseline 3, 6, and 12 months follow-up. As of the end of September 2023, the overall 3-month and 6-month survey completion rate is 85% (83% in SCP arm; 87% in PNP arm) and 83% (84% in SCP arm; 81% in PNP arm), respectively. As of the end of September 2023 the number of Veterans in the survey study with baseline surveys were n=978 with n=764 completing 3-month and n=657 6-month surveys.

For the updated primary analysis, we will supplement the lower than expected 3-month EHR completion rate with the PROMIS measures collected from the survey when the 3- or 6-month follow-up survey is in the 2-4 month follow-up window for the EHR sample. In all cases, the 3-month EHR PROMIS measures and the survey measures are collected by telephone. In the EHR sample, data is collected in templated notes by clinical personnel at the clinic or from our central delivery team. Survey sample data is collected by blinded study personnel.

For the EHR sample, of those through 4-months of the study (n=1540), baseline demographic characteristics, rates of Centers for Disease Control and Prevention (CDC) high impact chronic pain and mean PROMIS scores for those with completed 3 -month EHR follow-up compared to those missing EHR 3-months follow-up are shown in **Table 1**. Those missing 3-month follow-up are slightly younger, have higher rate of White race and slightly lower rates of CDC high impact chronic pain. Baseline PROMIS measures for pain interference, physical function and sleep are similar between those that completed 3 -month vs. missing 3-month outcome. As of the end of September 2023, 44% of n=1752 enrolled subjects are a part of the survey sample (**Table 2**). Baseline characteristics and PROMIS survey measure scores for the EHR and Survey sample are shown in **Table 3**. Overall the survey sample is slightly younger, similar gender and White race distribution, with somewhat lower Black race representation in Survey sample than EHR sample and similar mean baseline PROMIS scores for pain interference, physical function and sleep disturbance.

**Table 1:** Baseline demographic characteristics, CDC high impact chronic pain rate and PROMIS measures of EHR sample (n=1752) and Survey sample (n=978) for those completed EHR 3-month follow-up vs. missing 3-month EHR follow-up.

| **Baseline Characteristic/Measure** | **Completed 3-month**<br>**N=777** | **Missing 3-month**<br>**N=763** |
| --- | --- | --- |
| **Female, n(%)** | 107(13.8) | 99(13.0) |
| **Race, n(%)** |  |  |
| White | 450(57.9) | 480(62.9) |
| Black | 262(33.7) | 207(27.1) |
| Other | 14(1.8) | 20(2.6) |
| Missing | 51(6.7) | 56(7.3) |
| **Hispanic Ethnicity** | 27(3.5) | 44(5.8) |
| **Age, mean(SD)** | 53.54(15.3) | 51.56(15.9) |
| **CDC High Impact Chronic pain, n(%)** |  |  |
| Referral | 596(76.7) | 585(76.7) |
| Baseline | 514(66.2) | 491(64.4) |
| **PROMIS Pain Interference, mean(SD)** | 63.28(6.9) | 62.80(6.7) |
| **PROMIS Physical Function, mean(SD)** | 36.45(4.7) | 36.98(4.7) |
| **PROMIS Sleep, mean(SD)** | 58.23(8.2) | 58.45(8.6) |

**Table 2:** Distribution of baseline survey in EHR enrolled sample by treatment arm.

| ARM | In EHR Not in Survey | In EHR, In survey | Total EHR enrolled |
| --- | --- | --- | --- |
| Arm 1 | 604 | 380 | 984 |
|  | 61.3% | 38.6% |  |
| Arm 2 | 386 | 382 | 768 |
|  | 50.3% | 49.7% |  |
| Total | 990 | 762 | 1752 |
|  | 54.2% | 43.5% |  |

**Table 3:** Baseline characteristics and PROMIS measures of EHR sample (n=1752) and Survey sample (n=978)

|  | EHR Sample |  |  | Survey Sample |  |  |
| --- | --- | --- | --- | --- | --- | --- |
| Patient Characteristics (from trial enrollment) | Arm 1<br>(N=984) | Arm2<br>(N=768) | Total<br>(N=1752) | Arm 1<br>(N=482) | Arm2<br>(N=496) | Total<br>(N=978) |
| Age, mean (SD) | 52.6(15.9) | 53.2(15.3) | 52.8(15.6) | 52.1 (15.5) | 51.4 (15.2) | 51.7 (15.3) |
| Sex– Female, N (%) | 109(11.1) | 109(14.2) | 218(12.4) | 48 (10.0) | 65 (13.1) | 113 (11.5) |
| Race, N (%) |  |  |  |  |  |  |
| Black or African American | 223(22.7) | 286(37.2) | 509(29.1) | 90 (18.7) | 163 (32.9) | 254 (25.9) |
| White | 664(67.5) | 415(54.0) | 1079(61.6) | 317 (65.8) | 285 (57.5) | 602 (61.5) |
| Ethnicity, N (%) |  |  |  |  |  |  |
| Hispanic or Latino | 54(5.5) | 28(3.6) | 82(4.7) | 85 (8.7) | 49 (10.2) | 36 (7.3) |
| **PROMIS Measures** |  |  |  |  |  |  |
| Pain Interference, Mean (SD) N | 63.0 (6.54)<br>956 | 63.0 (7.20)<br>766 | 63.0 (6.84)<br>1722 | 63.1 (6.61)<br>467 | 63.3 (6.68)<br>474 | 63.2 (6.64)<br>941 |
| Physical Function, Mean (SD) N | 37.0 (5.12)<br>958 | 37.3 (5.68)<br>764 | 37.1 (5.37)<br>1722 | 37.9 (5.58)<br>476 | 38.2 (5.87)<br>492 | 38.1 (5.73)<br>968 |
| Sleep Disturbance Mean (SD) N | 58.5 (8.31)<br>956 | 58.1 (8.49)<br>766 | 58.3 (8.39)<br>1722 | 58.7 (7.76)<br>479 | 58.5 (7.82)<br>489 | 58.6 (7.79)<br>968 |

There are 176 Veterans (75 in SCP arm; 101 in PNP arm) with missing EHR 3 - month outcomes where at least 4-months have elapsed since their EHR baseline and have 3- or 6-month survey outcomes in the 2-4 month follow-up window (**Table 4**). Use of these data boosts the overall 3-month completion rate by 11% to 62% (increased to 63% for the SCP arm and 61% for the PNP arm). Baseline mean PROMIS scores by EHR-survey overlap are similar and shown in **Table 5**.

**Table 4:** For Veterans that have an EHR baseline and through 4 months of study (n=1540) as of 9/30/2023 the distribution of 3-month outcomes from EHR and 3- and 6-months surveys.

| ARM | IN EHR, Completed 3-month | IN HER, Completed 3-month with survey boost | In EHR, no 3-M EHR, no survey or no survey in window | In EHR, no 3M EHR, 3-month survey in window | In EHR, no 3M EHR, 6-month survey in window | Total |  |
| --- | --- | --- | --- | --- | --- | --- | --- |
| ARM 1 | 401 | 502 | 318 | 96 | 5 | 101 | 820 |
|  | 48.9% | 61.2% | 38.7% | 11.7% | 0.6% | 12.3% |  |
| ARM 2 | 376 | 451 | 269 | 68 | 7 | 75 | 720 |
|  | 52.2% | 62.6% | 37.4% | 9.4% | 1.0% | 10.4% |  |
| Total | 777 | 953 | 587 | 164 | 12 | 176 | 1540 |
|  | 50.5% | 61.8% | 38.1% | 10.6% | 0.8% | 11.4% |  |

**Table 5.** Baseline PROMIS scores by EHR – Survey Overlap.

| **EHR – Survey Overlap** | **EHR**<br>**Mean(SD)** | **Survey**<br>**Mean(SD)** |
| --- | --- | --- |
| **In EHR, In Survey (n=763)** |  |  |
| Pain Interference | 62.8(7.0) | 63.1(6.7) |
| Physical Function | 36.7(4.6) | 38.0(5.7) |
| Sleep | 58.7(8.2) | 58.3(7.8) |
| **In EHR, Not In Survey (n=989)** |  |  |
| Pain Interference | 62.8(7.0) | - |
| Physical Function | 36.7(5.0) | - |
| Sleep | 58.0(8.6) | - |
| **Not in EHR, In Survey (n=200)** |  |  |
| Pain Interference | - | 63.6(6.6) |
| Physical Function | - | 38.3(5.8) |
| Sleep | - | 59.9(7.8) |

We did not need to reevaluate power as our initial power calculations for 90% power included a range of ICC values and we had effect size differences for both a lower sample size for survey outcomes and EHR outcomes (**Table 6**). We can use the effect size difference for the survey outcomes, where n=42 at 3-months follow up as that would be a 60% attrition rate for the EHR sample – so the effect size differences we can detect for an ICC of 0.01 range from 0.30 – 0.37, for attrition rates ranging from 20% to 60% for the EHR Veterans. We used the sample size calculator from the NIH website (https://researchmethodsresources.nih.gov/) and **Table 7** shows the output from that calculator that yields a range of effect size differences for different numbers of clinics per arm as well as number of units per clinic needed at 3-months for an ICC of 0.01. Based on this table with our projected loss to follow-up adjusted based on using the survey data, we expect our loss to follow-up to be in the 35-40% range and with an ICC of 0.01 would give 90% power to detect effect size differences in the 0.32 -0.33 range.

**Table 6.** Power Table for AIM-BACK.

| Number of clinics per condition | Number of patients per clinic needed at post-intervention follow-up (Total N) | Df for baseline covariate adjustment | Minimum Detectable Difference (Effect size)<br>ICC=0.01 | Minimum Detectable Difference (Effect size)<br>ICC=0.02 | Minimum Detectable Difference (Effect size)<br>ICC=0.05 | Number needed at baseline with 20% attrition per clinic (Total Nb) |
| --- | --- | --- | --- | --- | --- | --- |
| 8 | 42 (N=672) | 5 | 0.37 | 0.42 | 0.54 | 53<br>(Nb=848) |
|  |  | 2 | 0.36 | 0.41 | 0.53 |  |
| 8 | 84<br>(N=1344) | 5 | 0.30 | 0.36 | 0.50 | 105<br>(Nb=1680) |
|  |  | 2 | 0.29 | 0.35 | 0.49 |  |

Table key: Effect sizes differences between arms based on the net difference between the two study conditions across two points in time (baseline and 3-months) at 3-months follow-up for 90% power, an alpha of 0.025, within-subject correlation = 0.50, standard deviation (SD) of outcome=1, adjustment for baseline covariates (max of 4 group level covariates based on constrained randomization that could take up between 2 and 5 degrees of freedom; we conservatively estimated no reduction in variance with inclusion of baseline covariates) and intraclass correlation coefficients (ICC) of 0.01, 0.02 and 0.05.

**Table 7.** Output table from NIH Website Power Calculator for an ICC=0.01.

| Standardized Detectable Difference |  |  |  |  |  |  |
| --- | --- | --- | --- | --- | --- | --- |
| Groups Per Condition |  |  |  |  |  |  |
|  |  | 7 | 8 | 9 | 10 | 11 |
|  | 42 | 0.4168 | 0.3727 | 0.3417 | 0.3182 | 0.2993 |
| Members | 63 | 0.3648 | 0.3262 | 0.2991 | 0.2785 | 0.2619 |
| Per | 84 | 0.3358 | 0.3002 | **0.2753** | 0.2563 | 0.2277 |
| Group | 105 | 0.3171 | 0.2835 | 0.2600 | 0.2420 | 0.2277 |
|  | 126 | 0.3040 | 0.2718 | 0.2492 | 0.2320 | 0.2183 |

Table Key: Effect sizes calculated using Research Methods Resources: National Institutes of Health. Available from: https://researchmethodsresources.nih.gov/

## Updated Analysis Plan

### Primary Analyses (Aim 1)

The primary outcomes are continuous and will be ascertained at the planned baseline and follow-up assessment (3 months) from administrative data collected from the EHR on Veterans presenting at participating clinics that are referred and enrolled in AIM - BACK. For Veterans in the EHR sample that do not have the 3-month EHR outcome data within the 3-month follow-up window but have also participated in the survey study, we will supplement the 3-month EHR with the PROMIS measures collected from the survey when the 3- or 6-month follow-up survey is in the 2-4 month follow-up window for the EHR sample. In all cases, the 3-month EHR PROMIS measures and the survey measures are being collected by telephone. In the EHR sample, data is collected in templated notes in the EHR by clinical personnel onsite or clinical personnel from our central delivery team. Survey sample data is being collected by blinded study personnel.

Changes in pain interference and/or physical function scores will be estimated and the primary hypotheses tested via hierarchical linear mixed-effects models, with patients nested within clinics and baseline and 3 month values in the response vector.[3] Hierarchical linear models are a flexible and powerful analytic tool for clustered longitudinal continuous outcomes. The fixed-effect portion of the model will have the form: *Y_ijk_ = β_0_ + β_1_*(followup) + β_2_*(followup*intervention)* for clinic *i*, patient *j,* at time *k*. Random effects (clinics and time by clinics) will be included in the model to account for clustering of patients within clinics as the clinics are the unit of randomization. Random effects will also be included to account for the within-patient correlation between repeated measures over time. We will determine the best-fitting random effects structure by fitting a variety of random coefficient models (e.g., random intercept only, random intercept and linear slope) and assessing model performance with the Akaike Information Criteria (AIC) model selection criteria.[4, 5] The predictors in the model will include a time effect and indicator variables for treatment interacting with the time effect. The intraclass correlation capturing the relationship of outcomes between patients seen at the same clinic is accounted for via the random effects for the clinics and time by clinics, which are assumed to be normally distributed. The model will be fit in the SAS procedure PROC MIXED using full likelihood approximation and the hypotheses will be tested by whether the estimated coefficient *β_2_* is positive and significantly different than 0 at the 0.025 level due to 2 primary outcome variables. We will include covariates used in the covariate constrained randomization[6] (5 potential variables; average pain scores, clinic location (main medical center/community clinic), number of participating primary care providers, average level of opioid exposure of LBP patients at clinic, and average age of LBP patients at clinic) in our primary model as well as a limited number of patient-level covariates that are readily available in the EHR (CDC pain, age, gender, race and a comorbidity measure).

The survey outcomes for the enrolled subset of patients will be collected at baseline, 3, 6, and 12 months and will include pain interference, function, intensity, catastrophizing, sleep, and depression. These are all longitudinal continuous outcomes and a hierarchical linear model similar to that described for the primary aim will be fit. We will fit random coefficient models as described above (e.g. random intercept only, random intercept and linear slope) and assess using AIC model selection criteria to determine the best model for the covariance structure. Similarly, we will determine the best model for the mean structure (e.g. linear, quadratic, dummy coding) as there are four outcome measurement occasions guided by descriptive plots and model fit assessed using AIC model selection criteria. Due to the timing of administration of baseline surveys – some baseline surveys may occur after the initial intervention contact - we will conduct a sensitivity analysis treating baseline surveys that occur after initial provider contact as occurring in the post-treatment period.

### Sensitivity Analyses

We are updating and expanding our sensitivity analysis. We originally proposed a sensitivity analysis fitting models that include all EHR follow-up measurements at their follow-up time (even those outside the 3-month follow-up window) and then set up a contrast to estimate the treatment arm difference at 3-months from this model. We plan to expand on this analysis and, for the EHR sample, include all measurement time points (EHR and/or survey); the number of measurement occasions per subject will vary, ranging from a minimum of 1 (EHR baseline only) to a maximum of 8 measurement occasions (**Table 8**), and be at unequal intervals. The zero-time point will be the EHR baseline, and survey time points and other EHR time points will be denoted as days from EHR baseline. As described above, we will fit random coefficient models and determine the best covariance and mean structure. A curve-fitting method (e.g., lowess, splines) will be used to estimate the average trends in outcomes and to inform the functional form of our mean trajectory model (e.g., linear or quadratic and other higher order terms). We will set up a contrast in this model to estimate the treatment difference at 3 -months, the primary time point, as well as contrasts to examine treatment differences at longer follow-up times (e.g., 6-months).

**Table 8:**
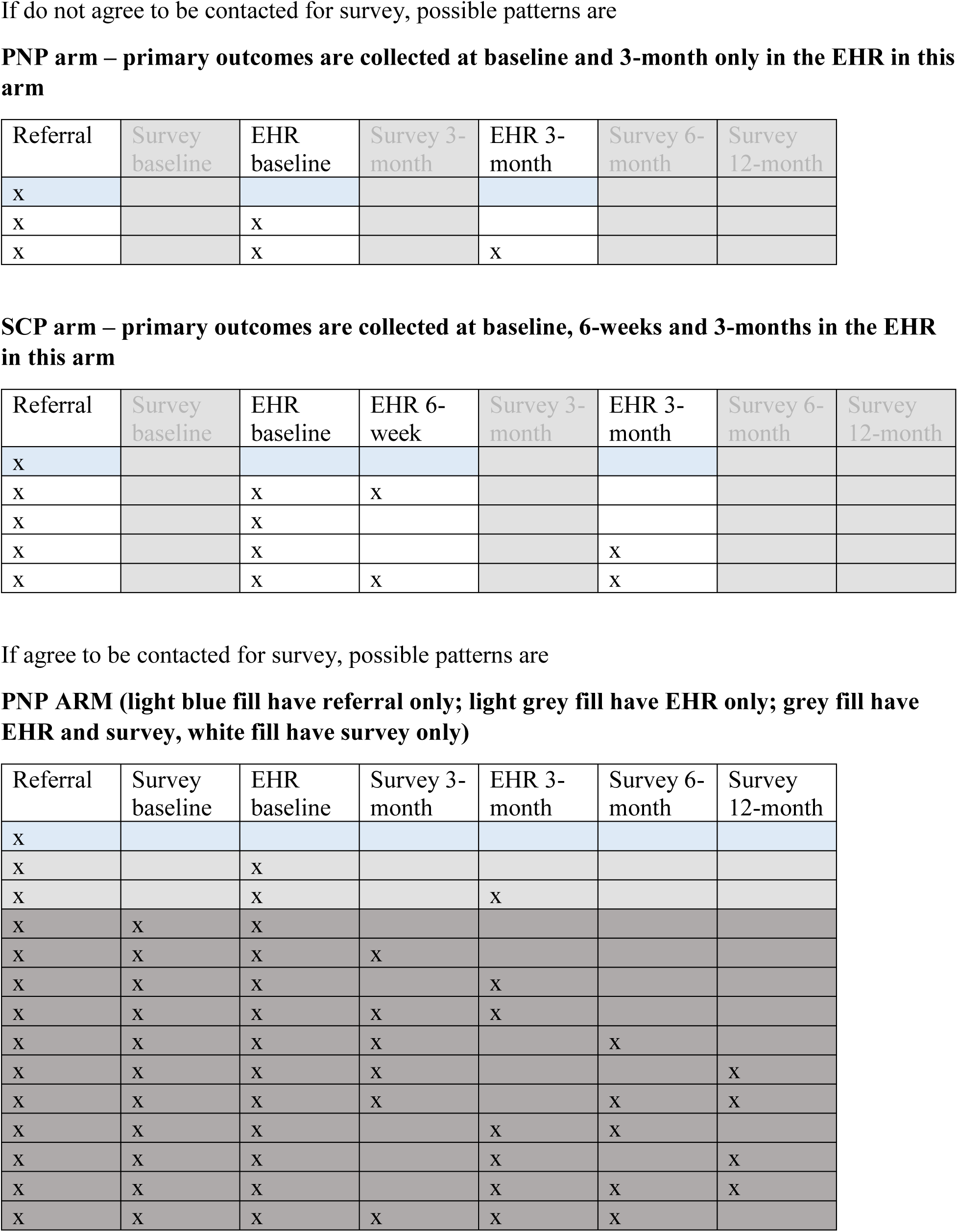
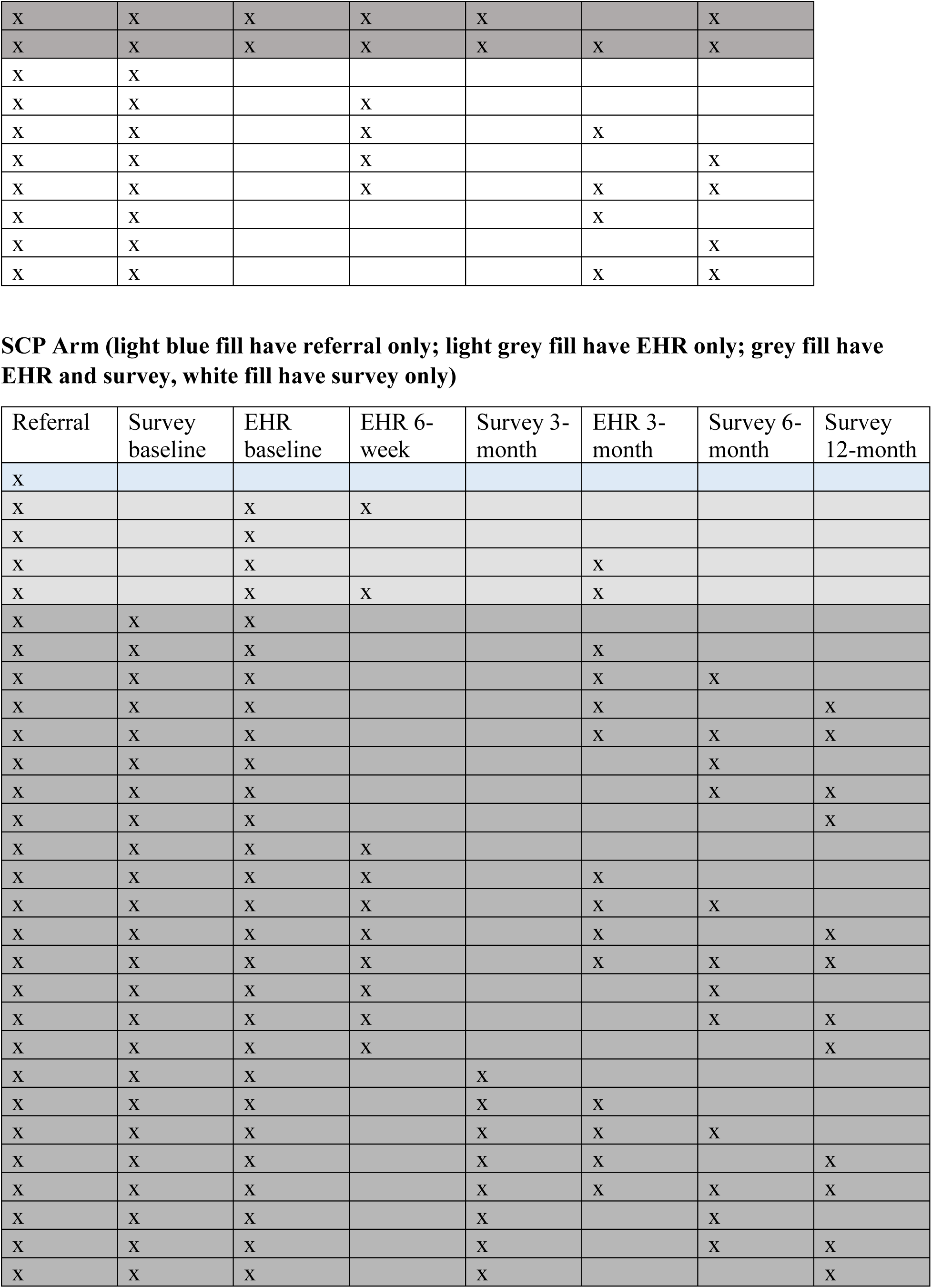
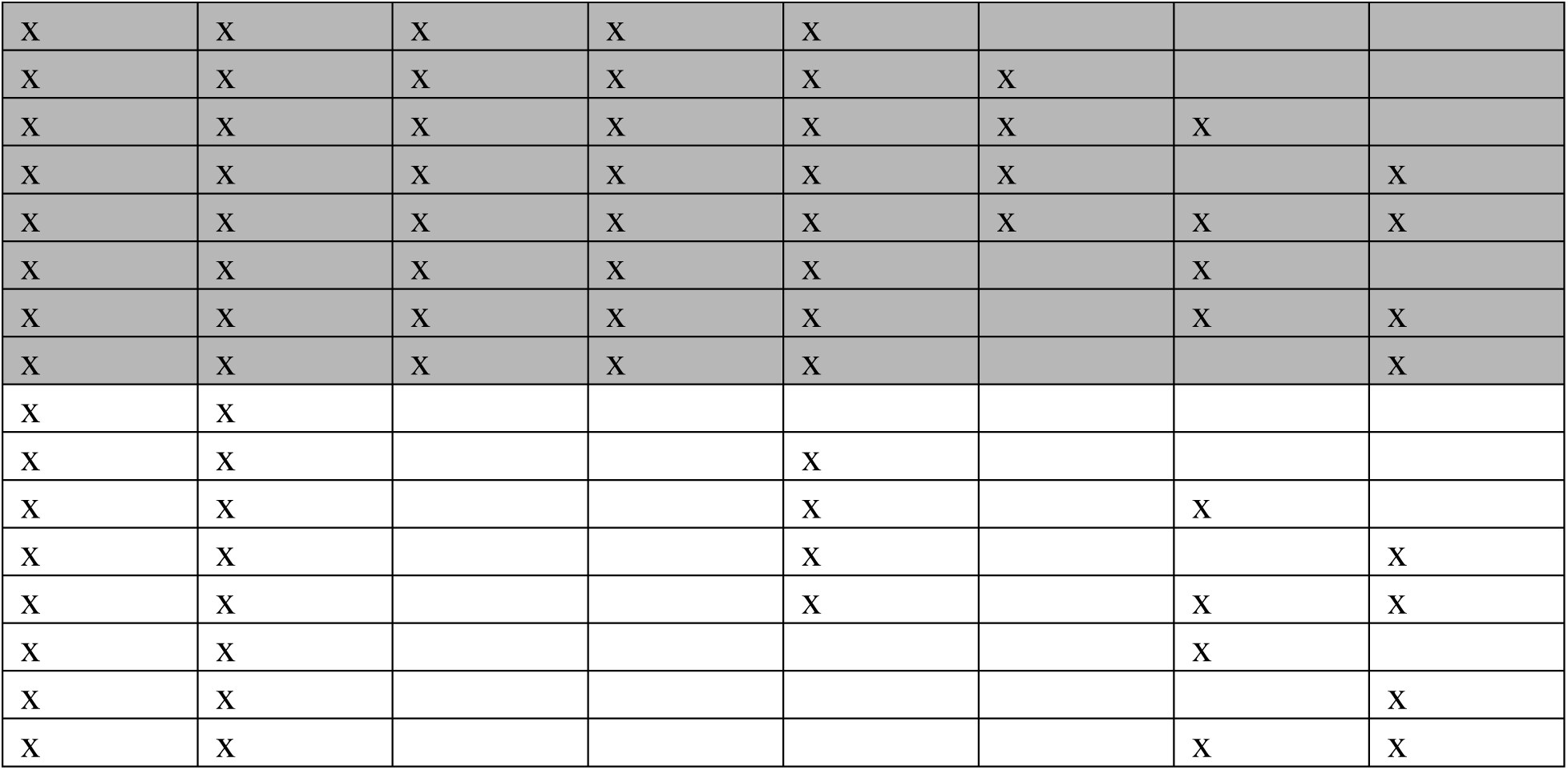
Data patterns for veterans referred by primary care providers at enrolled clinics to AIM-BACK pathway including baseline 3, 6 and 12-months survey.

### Missing Data

We do not anticipate much missing data in the main predictors of interest, intervention arm, or patient characteristics available in the EHR or assessed at baseline in survey outcomes. There may be missing values in the outcome measures due to dropout, death, a missed interim assessment or an assessment outside the 1 -month window for 3-month follow-up, or item non-response. However, hierarchical linear mixed models via maximum likelihood estimation, our main analysis technique for the primary outcomes, implicitly accommodate missingness at random (MAR).[3] Therefore, inferences will be valid even with differential dropout by intervention arm. We will thoroughly explore reasons for dropout and, depending upon the type and scope of missing data, we may explore the sensitivity of intervention effects to different missing data mechanisms (MAR vs. MNAR).[7] Our plan would be to conduct sensitivity analyses to evaluate the sensitivity of the assumption of MAR and/or MNAR on intervention effects. If our primary model included design variables only (treatment, stratification variables, clinic), this model would meet the MAR assumptions if missing data is related to previous outcome assessments or design variables included in the primary model. We can assess the sensitivity of the MAR assumption by conducting an analysis that includes auxiliary variables (either as additional variables in primary models or by conducting a multiple imputation including auxiliary variables). These auxiliary variables or other baseline characteristics for the large sample may be limited due to availability in the EHR and will explore additional variables including baseline opioid exposure and area depravation index. We are currently proposing to include age, sex, race, CDC pain and a comorbidity index in our primary analysis that may strengthen our MAR assumption. To explore the MAR assumption, outcomes will be multiply imputed using principled methods in SAS (via PROC MI or IVEware).[8] Multiple imputation provides a framework for being able to incorporate information from important auxiliary variables while still preserving a parsimonious main treatment effect model and is described as a significant advantage in recommendations from Panel on Handling Missing Data in Clinical Trials.[9] Note that if needed, we will utilize imputation methods that account for the multiple levels of correlation inherent in the clustered data structure. If we cannot justify the assumption of MAR, we will explore the sensitivity of intervention effects to the MNAR assumption; we will follow guidelines in Mallinckrodt[10] and Ratitch et al[7] for model selection and pattern mixture modeling.

In contrast to an EHR study where all measurements are determined by clinical practice and do not follow a data collection schedule, the AIM-Back program has a data collection schedule that has been implemented in the clinical process with outcome assessments documented in the EHR. Thus, standard missing data techniques for missing outcome measurements can be applied for those enrolled, as described above. However, because Veterans are referred to the AIM-BACK program by providers and must attend the first AIM-BACK visit to be enrolled, the potential for selection bias exists with a significant number of Veterans referred that are not enrolled and have no outcome data.

Through September 2023, we have approximately 2700 referrals with over 1700 Veterans enrolled in the program; approximately 70% of referrals led to enrollment in the AIM-BACK program with around 30% of referrals discontinued or cancelled across clinics. In monthly reports during the course of our study, we have been examining differences in CDC pain, age, gender, race, and ethnicity of Veterans referred and enrolled in the program to Veterans referred that do not enroll in the program. Overall to date, the patient demographics and CDC pain rates are similar between those enrolled and not enrolled. However, as we have no outcome data on those that do not enroll, in sensitivity analyses we will use inverse probability weighting methods (IPW) to adjust for this selection bias.[11] Probability weights for the probability of enrolling in the AIM-BACK program will be generated by fitting a logistic regression model to the binary enrollment indicator, with covariates for patient EHR demographics, and CDC pain at time of referral; we will explore whether adjustment for clinic is needed either as a fixed or random effect. We will adjust our primary model described above with probability weights and estimate treatment effects at 3-months.

## Conclusion

An updated plan for the primary analysis for the AIM-Back trial has been created and was approved by our monitoring partners on December 18, 2023. The need for an updated analysis plan was driven by the lower than projected follow-up rates for the primary outcome in the EHR observed during the AIM-Back trial. The updated plan does not change the primary outcome measures, but allows for clinical data to be supplemented by survey data that occurs within a 1 month timeframe around the 3 -month endpoint. Additionally, we present updated models for our sensitivity and missing data analyses. This updated analysis plan, along with the previously published protocol paper[1], are key source documents for evaluating the primary results of care pathway effectiveness from the AIM-Back trial.

## Data Availability

All data produced in the present work are contained in the manuscript

## Notes

### Competing Interest Statement

The authors have declared no competing interest.

### Clinical Protocols

https://pubmed.ncbi.nlm.nih.gov/33313728/

### Funding Statement

National Institutes of Health/National Center for Complementary and Integrative Health by UG3/UH3 grant AR009790

### Author Declarations

The Institutional Review Boards of Duke University and Durham VA Medical center gave ethical approval for this work

